# A systematic review of sample size estimation accuracy on power in malaria cluster randomised trials measuring epidemiological outcomes

**DOI:** 10.1101/2024.04.08.24305485

**Authors:** Joseph Biggs, Joseph D. Challenger, Joel Hellewell, Thomas S. Churcher, Jackie Cook

## Abstract

**Introduction:** Cluster randomised trials (CRTs) are the gold standard for measuring the community-wide impacts of malaria control tools. CRTs rely on well-defined sample size estimations to detect statistically significant effects of trialled interventions; however these are often predicted poorly by investigators. Here, we review the accuracy of predicted parameters used in sample size calculations for malaria CRTs with epidemiological outcomes.

**Methods:** We searched for published malaria CRTs using four online databases in March 2022. Eligible trials included those with malaria-specific epidemiological outcomes which randomised at least six geographical clusters to study arms. Predicted and observed sample size parameters were extracted by reviewers for each trial. Pair-wise correlation coefficients were calculated to assess the accuracy of predicted control-arm outcome estimates and desired relative reductions (effect sizes) between arms compared to what was observed. Among trials which retrospectively calculated an estimate of heterogeneity in cluster outcomes, we recalculated study power according to observed trial estimates.

**Results:** Of the 1889 records identified and screened, 108 articles were eligible and comprised of 71 malaria CRTs. Among trials that included sample size calculations (91.5%, 65/71), most estimated cluster heterogeneity using the coefficient of variation (k) (80%, 52/65) which were often predicted without using prior data (67.7%, 44/65). Predicted control-arm epidemiological outcomes correlated weakly with those observed, with 61.2% (19/31) of prevalence estimates overestimated. Among the minority of trials which retrospectively calculated cluster heterogeneity (20%, 13/65), empirical values contrasted with those used in sample size estimations and often compromised study power. Observed effect sizes were often smaller than had been predicted at the sample size stage (72.9%, 51/70) and were typically higher in the first, compared to the second, year of trials. Overall, effect sizes achieved by malaria interventions tested in trials decreased between 1995 and 2021.

**Conclusions:** Study findings reveal sample size parameters in malaria CRTs were often inaccurate and resulted in underpowered studies. Future trials must strive to obtain more representative epidemiological sample size inputs to ensure interventions against malaria are adequately evaluated.

**Registration:** This review is registered with PROSPERO (CRD42022315741).

## Introduction

Malaria is a parasitic disease that in 2022 was responsible for the deaths of 608,000 individuals worldwide; most of whom were children in Sub-Saharan Africa [1]. There are numerous, effective interventions that can be used to combat malaria transmission that are recommended by the World Health Organisation (WHO). To generate evidence for the recommendation of these tools, cluster randomised trials (CRTs) are conducted to demonstrate the community-wide impacts [2]. Historically, CRTs have been instrumental in demonstrating the mass effects of the first insecticide-treated bed nets (ITNs) [3-5], mass chemoprevention strategies [4, 6], and most recently, dual-action, long-lasting insecticide treated nets (LLINs) [7, 8]. Despite their necessity, CRTs are subject to major constraints. Trialling interventions over large geographical areas is costly, logistically challenging, and at the design stage, requires well-defined estimates of underlying transmission patterns in the study setting [9]. Consequently, in recent years, some malaria CRTs have reported being underpowered and have presented inconclusive findings [10-14].

Investigators power CRTs according to sample size estimations which account for cluster-level randomisation, where groups of people, as opposed to individuals, are randomised to receive interventions. This design can result in heterogeneity of outcomes between and within clusters owing to groups of individuals, such as households, schools, and geographical areas, sharing similar biological and socio-economic characteristics which introduces correlation in study outcomes [15, 16]. Consequently, cluster heterogeneity needs to be incorporated into sample size estimations, along with expected control arm transmission and effect size estimates (relative percentage reductions between arms), to compensate for the lower precision associated with this design. The between- and within-cluster heterogeneity can be measured using the coefficient of variation (k) or intra-cluster correlation coefficient (ICC), respectively, and heavily impacts trial size [15, 16]. Trialling new interventions in areas with missing or inadequate data results in investigators having to rely on judgement-based estimates for their sample size estimations which may be inaccurate.

Numerous reviews have evaluated sample size estimations in CRTs focused on cancer treatments [17], school-based interventions [18], oral health [19], residential care [20] and CRTs in general [21]. These reviews highlighted that despite trials mostly including sample size estimations, not all calculations accounted for cluster heterogeneity (73% [17], 78% [18], 71% [19] and 47% [20]). Two of these reviews further explored whether trials also included empirical measures of cluster heterogeneity and compared them to prior estimates [18, 21]. Both reviews highlighted trials rarely provided retrospective estimates of cluster heterogeneity (<40%), and among trials that did, large differences were identified between predicted and observed estimates. This suggests many trialists misclassified the true degree of cluster heterogeneity at the design stage. Finally, one review explored which trials stated their desired effect sizes and compared them to those observed [21]. They showed that 68% of desired effect sizes were overestimated. Interestingly, none of these reviews compared the outcome measures predicted and observed in the control arms of their included trials. This is crucial as misclassification of predicted effect size, cluster heterogeneity and control-arm outcome measurements all impact study power [15, 16, 22].

Malaria transmission is driven by numerous environmental and socio-economic factors including rainfall, temperature, vegetation cover, type of housing and provision of malaria interventions [23-26]. Consequently, transmission is often spatially and temporally variable across various geographical scales. This presents a challenge for malaria CRTs as heterogenous transmission in the community may result in spatial/temporal variability in malaria-specific outcomes between geographical clusters. Therefore, estimating the level of malaria transmission in the control arm and the degree of cluster heterogeneity for malaria CRT sample size estimations is difficult in the absence of baseline data.

In this review, our aim was to investigate the characteristics and quality of sample size estimations in malaria CRTs that used geographical clusters. Specifically, we explored whether study investigators accurately predicted sample size estimation parameters, including control-arm transmission, cluster heterogeneity, and desired effect sizes, according to observed measurements during trials. It is hoped results from this review will improve future study design and help ensure trialists are able to accurately detect impacts of interventions that are vital in the fight against malaria.

## Methods

### Search strategy and selection criteria

We conducted a systematic review of published malaria CRTs with epidemiological outcomes. In March 2022, we searched the database systems Pubmed, Web of Science, Embase and Cochrane reviews using truncated versions of the terms ‘malaria’ and ‘cluster randomised trial’ for trials published in English language. The bibliographies of identified reviews were additionally screened according to title and abstract. Search results were imported into the reference manager Endnote where digitally identified duplicates were removed. Manually identified duplicates were removed by two reviewers (JB & JH). Pre-determined eligibility criteria were used to screen identified articles based on title and abstract (JB & JH) while screening discordance was adjudicated by consensus (TC & JC). Identified studies were eligible for inclusion if they met the following criteria: the study was a CRT wherein at least six geographical clusters were randomised to intervention and control arms; the study measured malaria-specific epidemiological outcomes. Such outcomes include malaria prevalence or incidence according to microscopy, rapid diagnostic tests (RDTs), or molecular methods. Trials that only measured anaemia and all-cause mortality were excluded as these outcomes could be attributed to other conditions. Prior to study initiation, the review was registered in PROSPERO on 9 March 2022 (CRD42022315741).

### Data extraction

Two reviewers (JB & JH) independently extracted information from the final list of studies. Extraction discrepancies were resolved by consensus with TC and JC. Data for sample size estimations and empirical outcomes were extracted for all epidemiological outcomes measured at all trial timepoints. For each trial, we extracted data on overall trial design, randomisation method and type of intervention evaluated. For each sample size estimation, we extracted data on all assumptions outlined as well as those used to estimate cluster heterogeneity. To compare sample size assumptions to observed trial outcomes, where data was available, we extracted arm-aggregated malaria prevalence (cases/survey population) and/or incidence data (cases/person-years) by trial year and overall.

### Data analysis

For each sample size estimation where observed prevalence/incidence data were available, we calculated the relative reduction (effect size) between intervention arms for the duration of each study and stratified by year. The effect size was calculated according to equations *A* and *B* where subscript (1) and (2) represent the control and intervention arms, respectively, while (r) and (p) correspond to malaria incidence per person year and malaria prevalence, respectively. In this manner, the effect size represents the % relative reduction between the control and the intervention arm.

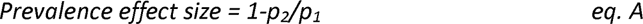

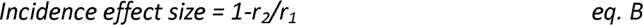

For sample size parameters including control-arm prevalence, incidence and effect size estimates, pair-wise Pearson’s correlation coefficients were used to calculate the strength of association between those estimated and observed for trials (Rho, p-value<0.05). To determine whether parameters were accurately estimated by investigators, we calculated the relative percentage difference between estimates and observed outputs. Parameters were classified as overestimated/underestimated if they exceeded a relative percentage difference of 10%. Regarding cluster heterogeneity estimates (k/ICC) provided in trials, we first investigated whether estimates based on pilot or baseline data differed to estimates based on no data. A pair-wise t-test was used to determine whether the mean value difference equalled zero. Among trials that reported cluster heterogeneity using observed trial data, we recalculated study power (%) according to the observed k/ICC and year 1 control arm prevalence/incidence but using the desired effect size quoted in the paper. The remaining sample size parameters used were identical to the original power calculations: desired effect size (%), cluster size, cluster number and significance level (%). Study power for CRTs was calculated according to methods described by Hayes and Moulton in [15]. All analyses were conducted in STATA (v.17, Texas, USA).

## Results

Our literature search yielded 1889 records from database searching and 145 records from the bibliographies of Cochrane reviews (Figure 1). Following the removal of duplicates, a total of 1302 records were screened after which 991 were excluded as they were not concerned with malaria CRTs. The remaining 311 records were assessed for eligibility resulting in 108 published articles being included in this study, which comprised of 71 malaria CRTs (Additional file 1). The review PRISMA 2020 checklist is included in additional file 2.

**Figure 1:**
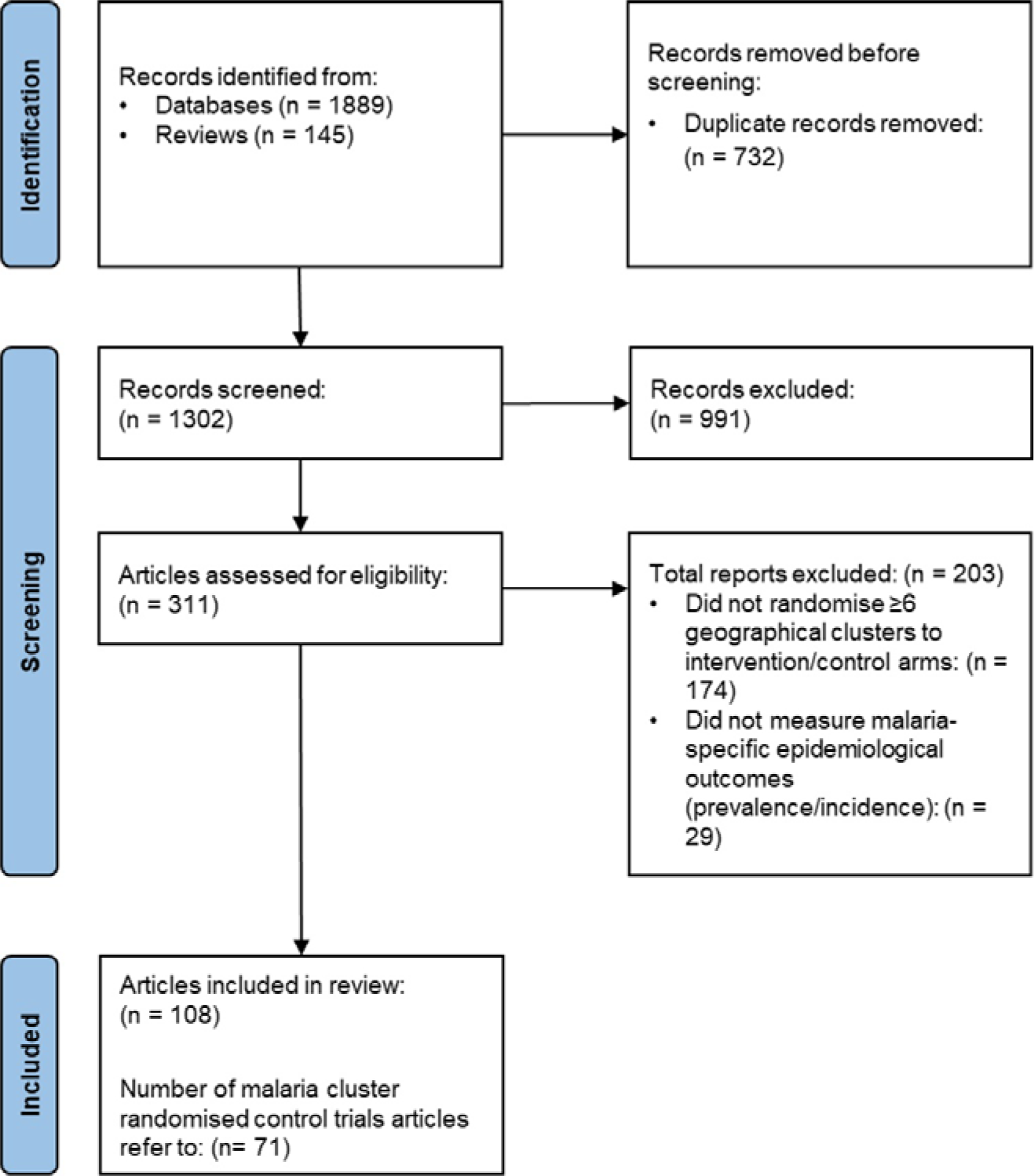
Study selection of included epidemiological malaria CRTs

The trial-level characteristics of the 71 malaria CRTs are shown in Table 1 and Figure 2A-G. Since 1995, the number of malaria-specific CRTs have increased in frequency and distribution across the world (Figure 2A&B). The 71 identified CRTs were conducted in a total of 78 countries, with study sites in Africa (N:53), Asia (N:21) and South America (N:4). 54.9% (39/71) of the trials evaluated vector control interventions, all measured *Plasmodium falciparum* outcomes while 26.8% (19/71) also measured *Plasmodium vivax* outcomes. Most trials adopted a parallel design (85.9%, 61/71) and consisted of two study arms (77.5%, 55/71). Concerning the cluster randomisation procedures, 39.4% (28/71) used simple randomisation to allocate clusters, 23.9% (17/71) implemented stratified randomisation, 22.5% (16/71) employed restricted randomisation, while 12.7% (9/71) randomised clusters within matched pairs. Among trials that randomised clusters through pair-matching or stratification, most restricted allocation based on a single criterion. For those that utilised restricted randomisation, most used between 3-4 restriction criteria (Figure 2F). The most common restriction criteria used for randomisation included cluster transmission intensity (prevalence or incidence), cluster size, location, and historical intervention coverage (Figure 2G). Regarding cluster design, 74.6% (53/71) adopted a basic cluster design, 14.1% (10/71) used a ‘fried egg’ design and 11.3% (8/71) reported ensuring a minimum buffer distance between clusters. Among trials that reported their minimum cluster buffer size, 72.7% (8/11) reported a minimum buffer size <2km while those who stated a minimum cluster separation reported a range between no separation and 3 km.

**Figure 2:**
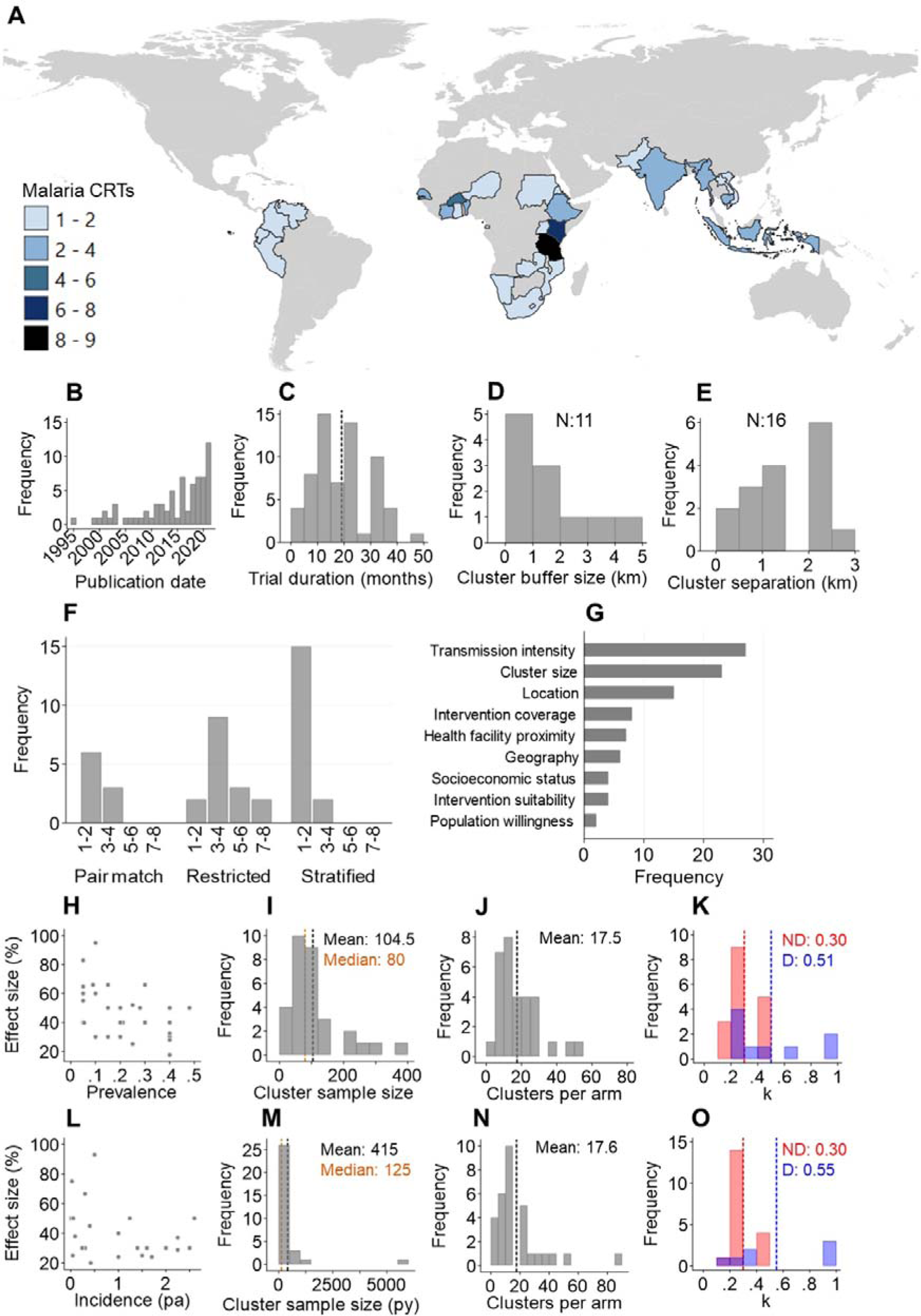
Characteristics of malaria CRTs identified in this review. **A**: Distribution of malaria CRTs. **B**: Annual frequency of malaria CRTs**C**. : Overall duration of malaria CRTs (dash line: mean). **D**: Size of buffers around study clusters. **E**: Minimum separation between study clusters. **F**: Number of restriction criteria used according to the type of trial randomisation. **G**: The most utilised restriction criteria in malaria CRTs. Population willingness refers to population acceptance of trialled interventions. Sample size assumptions used in trials with prevalence as the outcome measure: **H**: Predicted control-arm prevalence compared to desired effect size. **I**: The desired total number of individuals surveyed per cluster. **J**: Required number of clusters per arm. **K:** the predicted coefficient of variation (k) for prevalence stratified by whether values were estimating using prior or baseline data (D) or assumed using no data (ND). Vertical dash: mean. Sample size assumptions used in trials with incidence as the outcome measure: **L**: Predicted control-arm incidence per person compared to desired effect size (pa: per annum). **M**: The desired person-years per cluster. **N**: Required number of clusters per arm. **O:** the predicted coefficient of variation (k) for incidence stratified by whether values were based on prior or baseline data (D) or assumed using no data (ND). Vertical dash: mean.

**Table 1:** Overall characteristics of malaria CRTs identified in the systematic review (N:71)

| **Trial characteristics** | **n** | **%** |
| --- | --- | --- |
| **Intervention type** |  |  |
| Community engagement | 1 | 1.4 |
| Drug | 26 | 36.6 |
| Drug & Vector | 5 | 7.0 |
| Vector | 39 | 54.9 |
| **Primary outcome** |  |  |
| Incidence | 39 | 54.9 |
| Prevalence | 32 | 45.1 |
| **Malaria species** |  |  |
| *P. falciparum* | 52 | 73.2 |
| *P. falciparum* & *P. vivax* | 19 | 26.8 |
| **Design** |  |  |
| Parallel | 61 | 85.9 |
| Factorial | 4 | 5.6 |
| Crossover | 2 | 2.8 |
| Stepped wedge | 4 | 5.6 |
| **Randomisation method** |  |  |
| Block | 1 | 1.4 |
| Pair matched | 9 | 12.7 |
| Restricted | 16 | 22.5 |
| Simple | 28 | 39.4 |
| Stratified | 17 | 23.9 |
| **Number of study arms** |  |  |
| 2 | 55 | 77.5 |
| 3 | 9 | 12.7 |
| 4 | 7 | 9.9 |
| **Cluster type** |  |  |
| Basic | 53 | 74.6 |
| Buffered | 8 | 11.3 |
| Fried egg | 10 | 14.1 |
| **Level of analysis** |  |  |
| Cluster | 23 | 32.4 |
| Individual | 48 | 67.6 |
| **Formal sample size equation<sup>a</sup>** |  |  |
| Yes | 53 | 74.6 |
| No | 18 | 25.4 |
| **Reported being underpowered<sup>b</sup>** |  |  |
| Yes | 17 | 23.9 |
| No | 54 | 76.1 |

*b: reported at the end of the trial*

Among the included 71 CRTs, a total of 65 formal cluster sample size estimations were conducted that accounted for cluster heterogeneity by including a k, ICC or design effect component. Of these, 34/65 were based on incidence while 31/65 were based on prevalence (Table 2 & Figure 2H-O). The remaining trials either did not account for cluster heterogeneity or lacked any sample size justification. Over 90% of all sample size estimations were calculated to achieve at 80% power (59/65) at the 5% significance level (60/65). Concerning the epidemiological outcome measures in the control arm, most investigators predicted incidence using prior data (70.6%, 24/34) while most investigators predicted prevalence without using prior data (54.8%, 17/31). Regarding sample size estimations based on prevalence (N: 31), investigators estimated a range of prevalences in the control arm (mean: 0.21, range: 0.05 – 0.48) and desired effect sizes (mean: 47.1% range: 17.5 - 95%) which tended to be higher in low prevalence settings (Figure 2H). The average required cluster sample size was 104.5 individuals (Median 80; Figure 2I) and average required number of clusters per arm equalled 17.5 (Figure 2J). For sample size estimations based on incidence (N: 34), a range of incidence estimates in the control arm were estimated (range: 0.002 - 2.6 cases per person per annum (pa)). Desired effect sizes (%) were similarly higher in lower incidence settings (mean: 41.1%, range 20 - 93%) (Figure 2L). The average cluster size for incidence was 415 (median: 125) person years (Figure 2M) while the mean number of required clusters per arm was 17.6 (Figure 2N).

**Table 2:** Characteristics of sample size estimations used in malaria CRTs stratified by outcome measure: prevalence or incidence. There were a total of 65 sample size estimations, 34 based on incidence outcomes while 31 based on prevalence outcomes.

| Sample size characteristics | Overall<br>N:65 |  | Incidence<br>N:34 |  | Prevalence<br>N:31 |  |
| --- | --- | --- | --- | --- | --- | --- |
|  | n | % | n | % | n | % |
| Desired power (%) |  |  |  |  |  |  |
| 80-90 | 59 | 90.8 | 29 | 85.3 | 30 | 96.8 |
| 90-100 | 6 | 9.2 | 5 | 14.7 | 1 | 3.2 |
| **Significance level (%)** |  |  |  |  |  |  |
| 10 | 1 | 1.5 | 0 | 0.0 | 1 | 3.2 |
| 5 | 60 | 92.3 | 32 | 94.1 | 28 | 90.3 |
| 2.5 | 2 | 3.1 | 1 | 2.9 | 1 | 3.2 |
| 1.67 | 2 | 3.1 | 1 | 2.9 | 1 | 3.2 |
| **Cluster heterogeneity measure** |  |  |  |  |  |  |
| ICC | 10 | 15.4 | 6 | 17.6 | 4 | 12.9 |
| Design effect | 3 | 4.6 | 2 | 5.9 | 1 | 3.2 |
| k (CV) | 52 | 80.0 | 26 | 76.5 | 26 | 83.9 |
| **Method used to estimate cluster heterogeneity** |  |  |  |  |  |  |
| With data<sup>a</sup> | 21 | 32.3 | 9 | 26.5 | 12 | 38.7 |
| Without data | 44 | 67.7 | 25 | 73.5 | 19 | 61.3 |
| **Method used to estimate control-arm transmission** |  |  |  |  |  |  |
| With data<sup>a</sup> | 38 | 58.5 | 24 | 70.6 | 14 | 45.2 |
| Without data | 27 | 41.5 | 10 | 29.4 | 17 | 54.8 |
| **Retrospectively estimated cluster heterogeneity** |  |  |  |  |  |  |
| Yes | 13 | 20.0 | 5 | 14.7 | 8 | 25.8 |
| No | 52 | 80.0 | 29 | 85.3 | 23 | 74.2 |
| **Diagnostic used** |  |  |  |  |  |  |
| PCR | 10 | 15.4 | 3 | 8.8 | 7 | 22.6 |
| RDT | 29 | 44.6 | 17 | 50.0 | 12 | 38.7 |
| Microscopy | 19 | 29.2 | 10 | 29.4 | 9 | 29.0 |
| Mixed | 7 | 10.8 | 4 | 11.8 | 3 | 9.7 |
| **Age range tested (years)** |  |  |  |  |  |  |
| <5 | 15 | 23.1 | 8 | 23.5 | 7 | 22.6 |
| <10 | 14 | 21.5 | 9 | 26.5 | 8 | 25.8 |
| <15 | 7 | 10.8 | 10 | 29.4 | 9 | 29.0 |
| All ages | 29 | 44.6 | 11 | 32.4 | 10 | 32.3 |

*a: estimated using baseline or prior data*

The most common cluster heterogeneity measure used in malaria CRT sample size calculations was the coefficient of variation (k) (80% 52/65). 67.7% (44/65) of estimated cluster heterogeneity measures were estimated with no prior data while only 32.3% (21/65) were estimated using baseline or pilot study data. Lastly, only a minority of investigators retrospectively calculated cluster heterogeneity using trial data (20%, 13/65) (Table 2).

### Control arm transmission intensity assumptions

We explored how accurately epidemiological outcomes were predicted in the control arms of included trials (prevalence N: 31, incidence N: 34). Throughout each of the trials, predicted prevalence and incidence only moderately positively correlated with corresponding observed values (incidence r: 0.49, p-value<0.05, prevalence r: 0.39, p-value<0.05) (Figure 3A&B). Moreover, most predicted prevalence and incidence estimates were overestimated by more than 10% according to observed estimates (prevalence overestimation: 61.1% (19/31); incidence overestimation: 50% (17/34)) (Figure 3C&D). We also assessed whether using prior data was better for predicting subsequent prevalence/incidence. Results revealed no different between using prior data or not (Figure 3A&B). Lastly, we found predicted prevalence/incidence correlated significantly with those observed in first year of each trial yet not the second year (Figure 3E&F). These results demonstrate investigators tended to poorly predict control-arm prevalence/incidence, particularly in the second year and that prevalence was often overestimated in malaria CRTs.

**Figure 3:**
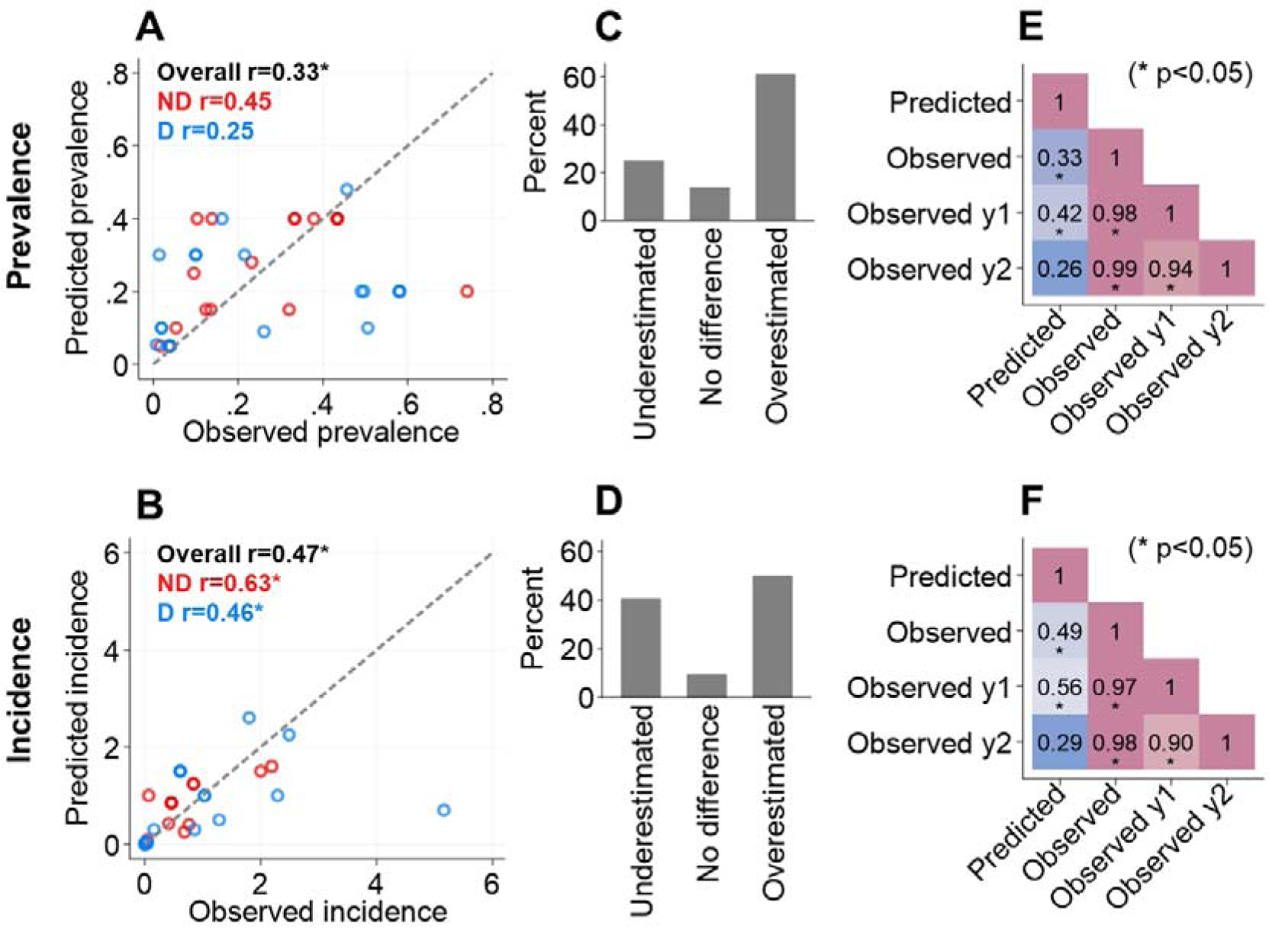
Accuracy of predicted versus observed prevalence and incidence outcomes in malaria CRT control arms. **A & B:** Correlation between the predicted and overall observed prevalence/incidence stratified by method used to predict estimates: using data (D; blue) using no data (ND; red) and combined (black). r: correlation coefficient. **C & D**: The percentage of predicted prevalence/incidence estimates that were underestimated (relative percentage difference <-10%), no difference (relative percentage difference -10% to 10%) or overestimated (relative percentage difference >10%) according to overall observed estimates. **E & F**: Correlation matrix comparing the predicted prevalence/incidence with estimates observed throughout the trial (observed), in year 1 (Observed y1) and in year 2 (Observed y2). *: r p-value<0.05.

### Cluster heterogeneity assumptions

Among trials that utilised the coefficient of variation (k) to account for cluster heterogeneity of incidence/prevalence in their sample size estimations (Table 2), a range of values were used (mean: 0.37, range: 0.1 - 1.0) (Figure 2K&O). Values of k predicted using prior data were, on average, statistically higher than those predicted with no prior data (no prior data mean k: 0.30; prior data mean k: 0.52, t-test p-value<0.05). This suggests k was likely underestimated in many trials. A small number of trials used the ICC to account for cluster heterogeneity, and they similarly had a large range (mean: 0.12, range: 0.006 – 0.40) (Additional file 3).

Among the trials that additionally calculated k/ICC at the end of the study using empirical data (20% 13/65), we explored whether predicted cluster heterogeneity estimates were accurate and then used the measured value for k/ICC to recalculate study power (Table 3). Empirical cluster heterogeneity estimates often differed to those used in sample size estimations with the majority underestimating k/ICC (61.5% 8/13). Recalculated power estimates, according to the stated desired effect, observed control arm transmission intensity and k/ICC, were often lower than originally planned for. The observed power for 7/11 trials was below 80%. For 4/11 trials, cluster heterogeneity was overestimated which resulted in them remaining suitably powered to detect their desired effect sizes. We were unable to replicate power calculations for two of the trials that provided retrospective k/ICC estimates [27, 28]. It should be noted it was not always stated which timepoint/subset of trial data was used to retrospectively calculate k/ICC.

**Table 3.**
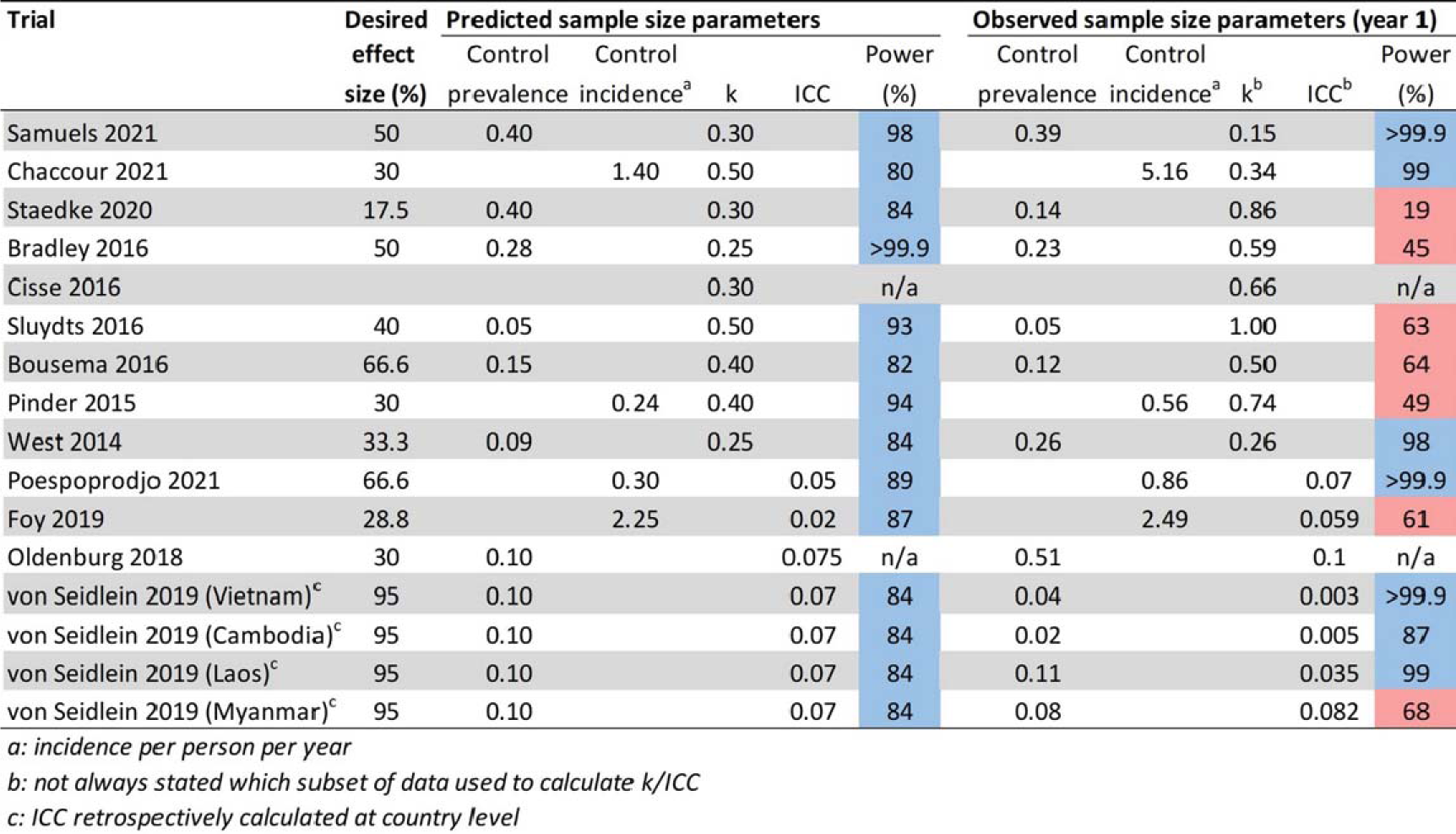
The study power (%) to detect desired effect sizes according to predicted (left) and observed (right) sample size parameters among trials the retrospectively calculated cluster heterogeneity. The predicted sample size parameters include the predicted control-arm prevalence/incidence and the k/ICC values stated in the article methods. The observed sample size parameters include the empirical control-arm prevalence/incidence and k/ICC values in the first year of the trials. First year data was utilised to estimate observed study power to account for temporal variations in transmission/cluster heterogeneity. The remaining sample size parameters including clusters per arm, cluster size and significance level were identical between the predicted and observed power calculations. Blue: study power >80%. Red: Study power <80%.

### Effect size assumptions

Among the 71 included malaria CRTs, a total of 70 desired effect size estimates were accompanied with empirical effect size estimates. We examined whether the desired effect sizes used in sample size estimations corresponded to observed effect sizes in malaria CRTs. We identified no evidence of a correlation between desired and observed effects sizes throughout the trial (r=0.21, p: 0.09) (Figure 4A). We also found that 72.7% (51/70) of desired effect sizes overestimated by more than a relative 10% difference (Figure 4B). We then explored factors that may have contributed to these findings. Firstly, among trials that were conducted for at least 2 years (N:36), we found a strong positive correlation between year 1 and year 2 observed effect sizes (r=0.49; p:0.003) (Figure 4C). Despite this, effect sizes were more often higher in the first, compared to the second, year of each trial (52.8% 19/36) (Figure 4C). Secondly, after comparing observed effect sizes to trial start date, a negative correlation was seen (r: -0.30, p<0.05), suggesting that effect sizes have been decreasing over time (Figure 4D).

**Figure 4:**
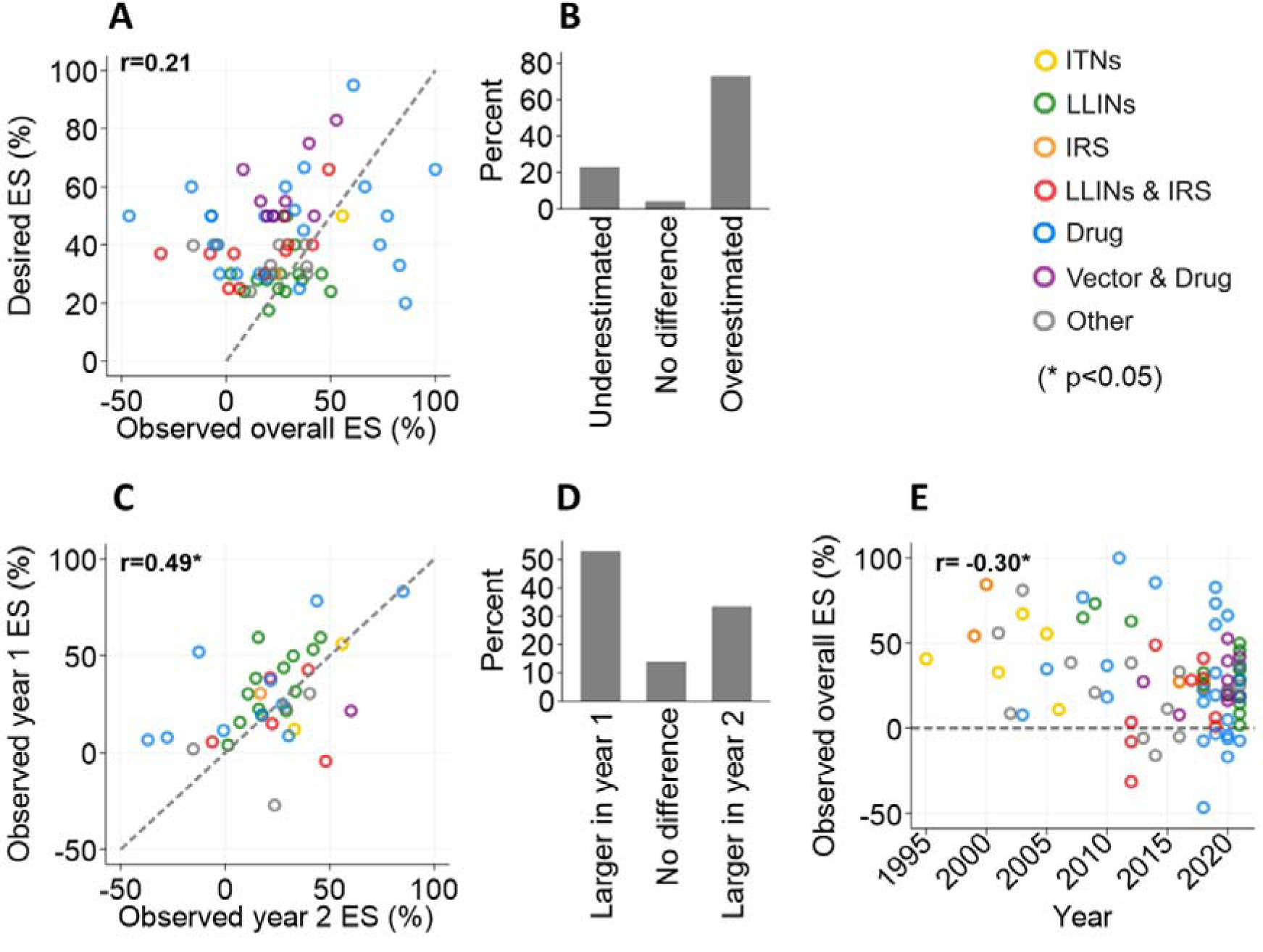
Accuracy of desired versus observed effect size (ES) estimates in malaria CRTs. **A**: Correlation between the desired and overall observed effect size by type of intervention. Diagonal dash: line of equality. **B**: The percentage of desired effect size estimates that were underestimated (relative percentage difference <-10%), no difference (relative percentage difference -10% to 10%) or overestimated (relative percentage difference >10%) according to overall observed effect size estimates. **C**: Correlation of observed effect size estimates by the 1 ^st^ and 2^nd^ year of the trial by type of intervention. Diagonal dash: line of equalit**D**y : The percentage of observed effect size estimates that were higher in the 1^st^ or 2^nd^ year of the trial (relative percentage difference>10%) or were no different (relative percentage difference<10%). E: **D**: Correlation between the overall observed effect size estimates versus the trial starting year by type of intervention. Horizontal dash: cut off line for positive effect size. Above this line epidemiological outcomes were lower in the intervention, compared to the control arms, of trials.

## Discussion

Results from this review reveal malaria CRTs, measuring epidemiological outcomes, often rely on poorly defined sample size assumptions which results in compromised study power. Well powered trials need accurate information on predicted transmission intensity in the control arm, the estimated heterogeneity of outcomes between or within clusters and desired effect size between study arms. We found that transmission intensity and effect sizes were often over-estimated, with measures of cluster heterogeneity commonly misclassified. To ensure future malaria CRTs are adequately powered to detect the impacts of control interventions, efforts need to be made to ensure sample size parameters are more reliably estimated at the trial design stage.

Our finding that most desired effect sizes in malaria CRTs were overestimated corresponds with results from a separate review of 300 non-disease specific CRTs which found 68% of trials experienced lower effect sizes than anticipated [21]. Authors speculated this over-estimation was likely attributed to trials being powered to detect minimally important differences between study arms and/or ineffective interventions being trialled. These are common challenges for malaria CRTs too. A 30% effect size was previously documented as the threshold for an intervention to have public health relevance and be cost-effective according to the WHO. These are likely highly ambitious targets for certain interventions [2], particularly when being compared to already effective interventions. Moreover, some trialists concluded their interventions were simply inadequate to curb malaria transmission [12, 29, 30]. It should be noted that malaria CRTs have also suggested other factors, unrelated to study power, that impeded their ability to demonstrate an impact including low coverage/adherence [31, 32], inappropriate study settings [33, 34] and poor quality control [35].

In this review we further explored patterns in observed effect sizes among malaria CRTs and revealed effect size estimates tended to be higher in the first compared to the second year of trials. This implies the adherence and community-wide impact of certain trialled interventions wane over time. For interventions such as bed nets, recent studies in Tanzania [36], Nigeria [37] and Nicaragua [38] have demonstrated net coverage, usage, physical integrity and insecticidal activity all decreased within a two-year period. Secondly, our results highlight observed effect sizes have, overall, decreased since the 1990s. This is likely a consequence of trialled interventions being increasingly layered over existing, widespread standard-of-care for malaria. Historically, control arms in malaria CRTs consisted of either no or substandard interventions including untreated nets and placebo treatments [3, 39, 40]. Recently however, control arms of trials typically include numerous, effective malaria interventions [12, 41-43] and sometimes only differ from intervention arms with regards to regiment [42, 44]. Together, these factors likely resulted in effect sizes being overstated.

Predicting malaria transmission intensity in the control arms of CRTs is challenging given the disease is so spatially and temporally heterogeneous [24, 25]. Here, we found no evidence that estimating control-arm transmission intensity using prior data provided a more accurate measures of prevalence or incidence in control arms at the end of trials. Moreover, estimated transmission intensity correlated with transmission more closely in the first, compared to the second, year of the trials. This is likely the consequence of environmental, seasonal, socio-economic, and behavioural changes that impact both human and mosquito populations [45, 46], and highlights the challenge in forecasting short term malaria transmission patterns [47-51].

In this review only 20% of included malaria CRTs retrospectively calculated cluster heterogeneity using trial data which resembles the previous review of 300 CRTs in general that found only 11% provided empirical cluster heterogeneity estimates [21]. Moreover, the finding that the majority of observed cluster heterogeneity measures differed to those inputted into sample size equations is concerning as study power is so heavily impacted by between/within cluster correlation [15]. Future malaria trials should adhere to CONSORT guidelines and provide empirical estimates of cluster heterogeneity to both inform future trials and assist reviewers in determining whether trials are adequately powered to detect their desired impact [21]. Moreover, given a recent secondary analysis of a malaria CRT in Tanzania demonstrated temporal changes in within-cluster cluster heterogeneity during the intervention period [52], providing empirical estimates of cluster heterogeneity at various timepoints during trials may further help decipher whether trials were adequately powered throughout the trial period [53]. As only a few malaria trials provided retrospective estimates of k/ICC, we were unable to investigate whether basing estimates on prior data or not assists in accurately characterising cluster heterogeneity. Moreover, it was not always clear which subset of the data k/ICC measures referred to and investigators may have been prompted to present observed cluster heterogeneity measures if the interventions failed to show impact. Consequently, characterising the true degree of cluster heterogeneity among a representative sample of malaria CRTs to inform future trials remains an imperative area of continued investigation.

## Conclusion

Results from this review demonstrate the accuracy of epidemiological inputs in malaria CRT sample/power size calculations require improvement. By simply reporting empirical cluster heterogeneity measures alongside published results, in line with CONSORT guidelines, future trials may be better informed to estimate suitable sample sizes. Determining trial transmission intensity and heterogeneity in the control arm remains a larger challenge given the sporadic nature of malaria transmission. Without more representative sample size parameters, future CRTs are at risk of being underpowered to detect the impacts of vital, novel control tools against malaria.

## Data Availability

All data produced in the present work are contained in the manuscript

## List of abbreviations

CRT: Cluster randomised trial
ES: Effect size
ICC: Intracluster correlation coefficient
IRS: Indoor residual spraying
ITN: Insecticide-treated bed nets
k: coefficient of variation
LLIN: Long-lasting insecticidal nets
pa: Per annum
PCR: Polymerase chain reaction
py: Person-year
RDT: Rapid diagnostic test
WHO: World Health Organisation

## Declarations

### Ethics approval and consent to participate

Not applicable

### Consent for publication

Not applicable

### Availability of data and materials

The datasets generated and/or analysed during the current study are available from the corresponding author on reasonable request.

### Authors’ contributions

Study conception: JC and TC. Literature searching and extraction: JB and JH. Data analysis: JB and JDC. Supervision: JC & TC. Manuscript preparation: JB. Manuscript editing and review: JB, JDC, TC and JC. The authors read and approved the final manuscript.

## Acknowledgements

Not applicable

