## Additional file 1 for "A systematic review of sample size estimation accuracy on power in malaria cluster randomised trials measuring epidemiological outcomes": Additional file 1.docx

**Additional file 1**: List of malaria cluster randomised control trials included in this systematic review.

| **Trial ID** | **Authors** | **Date** | **Journal** | **Trial registration number** |
| --- | --- | --- | --- | --- |
| 1 | Poespoprodjo JR, Burdam FH, Candrawati F | 2021 | Lancet Infectious Diseases | NCT02787070 |
| 2 | Bath D, Cook J, Govere J | 2021 | Lancet | NCT02556242 |
| 3 | Vilakati S, Mngadi N | 2021 | BMJ | NCT02315690 |
| 4 | Sternberg ED, Cook J, ahiua Alou LP | 2021 | Lancet | ISRCTN18145556 |
| 5 | A. M. Samuels N. A. Odero W. Odongo K | 2021 | Clinical Infectious DIseases | NCT02987270 |
| 6 | Chaccour C, Zulliger R, Wagman J | 2021 | Malaria J | NCT02910934 |
| 7 | Minakawa N, Kongere JO, Sonye GO | 2021 | Am J Trop Med Hyg | UMIN000019971 |
| 8 | McLean ARD, Indrasuta C, Khant ZS | 2021 | Lancet Infectious Diseases | NCT01872702 |
| 9 | Dabira ED, Soumare HM, Conteh B | 2021 | Lancet Infectious Diseases | NCT03576313 |
| 10 | Okebe J, Dabira E, Jaiteh F | 2021 | Malaria J | NCT02878200 |
| 11 | Accrombessi M, Cook J, Ngufor C | 2021 | BMC Infectious Diseases | NCT03931473 |
| 12 | Mosha J, Kulkarni MA, Messenger L | 2021 | Lancet | NCT03554616 |
| 13 | Hsiang MS, Ntuke H, Roberts KW | 2020 | Lancet | NCT02610400 |
| 14 | Staedke SG, Gonahasa S, Dorsey G | 2020 | The Lancet | ISRCTN17516395 |
| 15 | Syafruddin D, Asih PBS, Rozi IE | 2020 | Am J Trop Med Hyg | NCT02294188 |
| 16 | Agius PA, Cutts JC, Oo WH | 2020 | PLOS Medicine | ACTRN12616001434482 |
| 17 | Eisele TP, Bennett A, Silumbe K | 2020 | Am J Trop Med Hyg | NCT02329301 |
| 18 | Desai MR, Samuels AM, Odongo W | 2020 | Am J Trop Med Hyg | NCT02987270 |
| 19 | Kone S, Utzinger J, Probst-Hensch N | 2020 | BMC Public Health | NCT04250428 |
| 20 | Arzika AM, Maliki R, Boubacar N | 2019 | PLOS Medicine | NCT02048007 |
| 21 | COSMIC Consortium | 2019 | Clinical Infectious Diseases | NCT01941264 |
| 22 | Foy BD, Alout H, Seaman JA | 2019 | The Lancet | NCT02509481 |
| 23 | Loha E, Deressa W, Gari T | 2019 | Malaria J | PACTR201411000882128 |
| 24 | Ndiaye JLA, Ndiaye Y, Ba MS | 2019 | PLOS Medicine | NCT01449045 |
| 25 | Lover AA, Dantzer E, Hocini S | 2019 | Gates open research | NCT03783299 |
| 26 | von Seidlein L, Peto TJ, Landier J | 2019 | PLOS Medicine | NCT01872702 |
| 27 | Oldenburg CE, Amza A, Kadri B | 2018 | Pediatr Infect Dis J | NCT00792922 |
| 28 | Morris U, Msellem M, Mkali H | 2018 | BMC Medicine | NCT02721186 |
| 29 | Protopopoff N, Mosha JF, Lukole E | 2018 | The Lancet | NCT02288637 |
| 30 | Tiono AB, Ouédraogo A, Ouattara D | 2018 | The Lancet | ISRCTN21853394 |
| 31 | Sutanto I, Kosasih A, Elyazar IRF | 2018 | Clinical Infectious Diseases | NCT01878357 |
| 32 | Manning J, Lon C, Sring M | 2018 | BMC Trials | NCT02653898 |
| 33 | Kafy HT, Ismail BA, Mnzava AP | 2017 | PNAS | NCT01713517 |
| 34 | Bridges DJ, Miller JM, Chalwe V | 2017 | BMC Trials | NCT02654912 |
| 35 | Bradley J, Hergott D, Garcia G | 2016 | Malaria J |  |
| 36 | Cissé B, Hadj Ba E, Sokhna C | 2016 | PLOS Medicine | NCT00712374 |
| 37 | Sluydts V, Durnez L, Heng S | 2016 | The Lancet Infectious Diseases | NCT01663831 |
| 38 | Homan T, Hiscox A, Mweresa CK | 2016 | The lancet | NTR3496 |
| 39 | Bousema T, Stresman G, Baidjoe AY | 2016 | PLOS Medicine | NCT01575613 |
| 40 | Mtove G, Mugasa JP, Messenger LA | 2016 | BMC Public Health | NCT02533336 |
| 41 | Zhou G, Wiseman V, Atieli H | 2016 | BMC Trials | NCT02392832 |
| 42 | Pinder M, Jawara M, Jarju LBS | 2015 | The Lancet | ISRCTN01738840 |
| 43 | Sangoro O, Turner E | 2014 | Malaria J | ISRCTN92202008 |
| 44 | Deressa W, Yihdego YY, Kebede Z | 2014 | BMC Parasites and Vectors | NCT01160809 |
| 45 | Tine RCK, Ndour CT, Faye B | 2014 | Trans R Soc Trop Med Hyg | PACTR201305000551876 |
| 46 | Kramer RA, Mboera LEG, Senkoro K | 2014 | Int J Environ Res Public Health |  |
| 47 | West PA, Protopopoff N, Wright A | 2014 | PLOS Medicine | NCT01697852 |
| 48 | Tiono AB, Ouédraogo A, Ouattara D | 2013 | Malaria J | NCT01256658 |
| 49 | Smithuis FM, Kyaw Kyaw M, Phe UO | 2013 | Malaria J |  |
| 50 | Bhatt RM, Sharma SN, Uragayala S | 2012 | Malaria J |  |
| 51 | Corbel V, Akogbeto M, Damien GB | 2012 | The Lancet Infectious Diseases | ISRCTN07404145 |
| 52 | Deribew A, Birhanu Z, Sena L | 2012 | Malaria J | ACTRN12610000035022 |
| 53 | Shekalaghe SA, Drakeley C, van den Bosch S | 2011 | Malaria J | NCT00509015 |
| 54 | Keating J, Locatelli A, Gebremichael A | 2011 | Acta Tropica |  |
| 55 | Tagbor H, Cairns M, Nakwa E | 2011 | Trop Med Int Health |  |
| 56 | Schellenburg JRM, Shirima K, Maokola W | 2010 | Am J Trop Med Hyg |  |
| 57 | Sharma SK, Tyagi PK, Upadhayay | 2009 | Acta Tropica |  |
| 58 | Thang ND, Erhart A, Speybroeck N | 2009 | PLOS ONE | NCT00853281 |
| 59 | Sahu SS, Vijayakumar M, Kalyanasundaram M | 2008 | Indian J Med Res |  |
| 60 | Magris M, Rubio-Palis Y, Alexander N | 2007 | Trop Med Int Health |  |
| 61 | Sochantha T, Hewitt S, Nguon C | 2006 | Trop Med Int Health |  |
| 62 | Henry MC, Assi SB, Rogier C | 2005 | Am J Trop Med |  |
| 63 | Kuile FOT, Terlouw DJ, Kariuki SK | 2003 | Am J Trop Med Hyg |  |
| 64 | Von Seidlein L, Walraven G, Miligan P | 2003 | Trans R Soc Trop Med Hyg |  |
| 65 | Macintyre K, Sosler S, Letipila F | 2003 | Int. J Epi |  |
| 66 | Habluetzel A, Cuzin N, Diallo DA | 2002 | Trop Med Int Health |  |
| 67 | Rowland M, Durrani N, Kenward M | 2001 | Lancet |  |
| 68 | Mnzava AE, Sharp BL, Mthembu DJ | 2001 | S Afr Med J |  |
| 69 | Rowland M, Mahmood P, Iqbal J | 2000 | Trop Med Int Health |  |
| 70 | Misra SP, Webber R, Lines J | 1999 | Trans R Soc Trop Med Hyg |  |
| 71 | Kroeger A, Mancheno M, Alacorn J | 1995 | Am J Trop Med Hyg |  |
