## Additional file 3 for "A systematic review of sample size estimation accuracy on power in malaria cluster randomised trials measuring epidemiological outcomes": Additional_File_3.docx

**Additional file 3**: Intra-cluster correlation coefficients (ICC) used in malaria CRT sample size calculations included in this review.

| **Trial** | **Outcome** | **ICC** | **Method used to estimate ICC** |
| --- | --- | --- | --- |
| Agius 2020 | Incidence | 0.15 | Without data |
| Foy 2019 | Incidence | 0.02 | Without data |
| Homan 2016 | Incidence | 0.006 | With data |
| Manning 2018 | Incidence | 0.4 | Without data |
| Poespoprodjo 2021 | Incidence | 0.05 | Without data |
| Arzika 2019 | Prevalence | 0.056 | With data |
| Kone 2020 | Prevalence | 0.25 | With data |
| Minkawa 2021 | Prevalence | 0.053 | With data |
| Oldenburg 2018 | Prevalence | 0.075 | Without data |
| Von Seidlein 2019 | Prevalence | 0.07 | Without data |
